# Factors influencing the COVID-19 daily deaths peak across European countries

**DOI:** 10.1101/2020.11.04.20225656

**Authors:** Katarzyna Jabłońska, Samuel Aballéa, Mondher Toumi

## Abstract

**OBJECTIVES:** The purpose of this study was to determine predictors of the height of COVID-19 daily deaths peak and time to the peak, in order to explain their variability across European countries.

**STUDY DESIGN:** For 34 European countries, publicly available data were collected on daily numbers of COVID-19 deaths, population size, healthcare capacity, government restrictions and their timing, tourism and change in mobility during the pandemic.

**METHODS:** Univariate and multivariate generalised linear models using different selection algorithms (forward, backward, stepwise and genetic algorithm) were analysed with height of COVID-19 daily deaths peak and time to the peak as dependent variables.

**RESULTS:** The proportion of the population living in urban areas, mobility at the day of first reported death and number of infections when borders were closed were assessed as significant predictors of the height of COVID-19 daily deaths peak. Testing the model with variety of selection algorithms provided consistent results. Total hospital bed capacity, population size, number of foreign travellers and day of border closure, were found as significant predictors of time to COVID-19 daily deaths peak.

**CONCLUSIONS:** Our analysis demonstrated that countries with higher proportions of the population living in urban areas, with lower reduction in mobility at the beginning of the pandemic, and countries which closed borders having more infected people experienced higher peak of COVID-19 deaths. Greater bed capacity, bigger population size and later border closure could result in delaying time to reach the deaths peak, whereas a high number of foreign travellers could accelerate it.

## Introduction

The coronavirus infectious disease 2019 (COVID-19) outbreak was announced as a pandemic by the World Health Organization (WHO) on 11 March 2020.^1^ By the end of March, Europe exceeded Asia and became the region experiencing the highest percentage mortality from the virus across the world,^2^ until 10 June 2020, when the rapidly growing COVID-19 mortality rate in the Americas exceeded all other continents.^3^ According to the WHO report from 5 July 2020, 38% of the global mortality due to COVID-19 was from Europe.^4^

Since incidence and mortality rates varied between countries, numerous studies have recently been published investigating factors associated with COVID-19 infection and death rate across countries. A variety of potential predictors have been assessed in the literature, such as country-specific demographic characteristics, economic and social indicators,^5–8^ mobility scores and social-distancing measures,^9, 10^ ecological and environmental perspectives,^8, 11^ as well as health characteristics and comorbidities that increase peoples’ vulnerability.^12^ To assess the relationship between covariates and COVID-19 incidence or mortality, the most common approach used by authors was to analyse multivariate regression models of daily or total number of infections or deaths up to a given time point as outcomes.

In this study, we used data on numbers of deaths, not infections, since the former has much higher degree of reliability than the latter, being better monitored and less dependent on the number of tests done. Since all European countries seem to already reach the peak of deaths by 3 June 2020 from the first wave of COVID-19 disease, our idea was to use height of daily deaths peak as a primary outcome and time to the peak as a secondary outcome.

To the best of our knowledge, this perspective has not been investigated so far. When raw and cumulative daily numbers of infections and deaths are subject to deviations from between-country differences in reporting and depend on date up to which the analysis is performed, maximum number of daily deaths can be assessed as an interesting new indicator of disease mortality magnitude.

This study aims to detect significant drivers of COVID-19 mortality with the use of multivariate generalised linear models (GLM) and distinct selection algorithms, to explain the variability of height of and time to the deaths peak among European countries. This will enable us to draw conclusions about how governments and societies can improve the future response on similar pandemic or global life-threatening situations.

## Methods

### Data collection

A total of 34 European countries were included in the analysis. The height of COVID-19 daily deaths peak was the primary outcome of interest, defined as maximum daily reported number of people who died due to COVID-19 per country up to 3 June 2020 divided by the number of inhabitants.

The secondary outcome of interest was time to COVID-19 daily deaths peak, defined as number of days from the day when the first death was reported in a country, up to the day of reaching the COVID-19 daily deaths peak.

A set of explanatory variables used to predict outcomes consisted of the following:

- Healthcare capacity:
  - All beds capacity (number of hospital beds)
  - Intensive care unit (ICU) capacity (number of ICU)
  - Number of tests conducted up to the peak
- Government restrictions and associated factors:
  - ‘Stay at home’ order date
  - Educational facilities closure date
  - Gathering restriction date
  - Businesses closure date
  - Border closure date
  - Total number of COVID-19 infected people when borders were closed
  - Total number of COVID-19 deaths when borders were closed
- Indicators of the population size:
  - Country population size (January 2020)
  - Percentage of population living in urban areas
  - Percentage of population living in metropolitan cities with more than 1 million inhabitants
- Median age
- Tourism:
  - Number of travellers that arrived at airports in 2018
  - Number of foreign tourists in 2018 (any touristic accommodation)
- Mean mobility score at the day of first reported COVID-19 death, calculated across mobility scores at:
  - Retail and recreation places
  - Workplaces
  - Transit stations.

All dates were considered as number of days after the day of first reported COVID-19 death in each country. Variables indicating healthcare capacity and tourism were considered in relation to the population size of a given country (per inhabitant or 1 million inhabitants).

#### Data sources

Data on COVID-19 deaths, infections, number of tests, beds capacity, government restrictions, population size and urban population size were taken from Institute for Health Metrics and Evaluation (IHME).^13^ Missing dates of government restrictions, if officially issued, were found on Wikipedia.^14^

Mobility scores were uploaded from Google COVID-19 Community Mobility Reports.^15^ Scores were reported as percentage changes from a usual mobility calculated before the pandemic in Europe, between January and February 2020. Scores were subject to oscillations due to daily reporting and presence of weekends and holidays; therefore, we used smoothed scores produced with a nonparametric technique that uses local weighted regression and fits a smooth curve through points in a scatter plot, called *loess regression*.^16^

Data on number of passengers arrived at airports, tourism and population living in metropolitan areas were downloaded from Eurostat^17^ and other sources.^18–21^

#### Missing data and imputation

If government restrictions were not officially set, the date was imputed with date of the peak in each country. Missing data on mobility for Cyprus and Iceland were imputed with average scores of remaining countries.

### Statistical analysis

Descriptive statistics on outcomes and explanatory variables were produced. Since the distribution of deaths peak height per population size was right-skewed, a logarithmic transformation was applied.

Factors influencing the height of deaths peak were analysed using univariate and multivariate GLM with a normal distribution function and logarithmic link function. The base case analysis was performed using data available for 34 countries and explanatory variables with p-value <0.1 in univariate GLM models. To avoid multicollinearity between independent variables, Pearson correlations were investigated to detect highly correlated pairs of variables. The correlation was assessed as high if the absolute value exceeded 0.7 and as moderate if the absolute value was in a range 0.5–0.7.^22^

Due to relatively low sample size, a risk of bias could appear for models with too high number of covariates. Therefore, sensitivity analyses were performed using variety of selection algorithms: stepwise, backward, forward and the genetic algorithm, to limit the number of covariates, increase model precision and improve model fit. A criterion of having p-value lower than 0.1 was applied for each variable to stay in a model (for backward and stepwise) and to enter a model (forward and stepwise). For the genetic algorithm, the best model was fitted based on the value of Akaike’s Information Criterion (AIC) corrected for small sample sizes (AICC). Models with only main effects were considered. Additional sensitivity analysis was performed removing countries for which imputation of government restriction dates was needed.

Similar methods were used to analyse time to deaths peak but without any transformation since it seemed to follow a normal distribution. GLM models with a normal distribution function and identity link function were analysed.

Given a country-level analysis, Moran’s I and Geary’s C statistics ^23, 24^ were produced to check the spatial autocorrelation in values of outcomes and to determine if it should be considered in models.

For all analyses, a p-value lower than 0.05 was considered as statistically significant. For each GLM model, fit statistics such as AIC and AICC were produced with lower values indicating better fit.

Analyses were performed using SAS 9.4 software. R 3.6.2 was used to apply the genetic algorithm with the package *glmulti*.^25^

## Results

### Descriptive analysis

Characteristics of countries included in the study are presented in Table 1, and a histogram of the height of COVID-19 deaths peak is depicted in Figure 1. The graph presenting deaths peak height by country is provided in Figure 2.

**Table 1.** Characteristics of countries included in the analysis.

| Variable | N | Mean | SD | Median | Lower quartile | Upper quartile | Min | Max |
| --- | --- | --- | --- | --- | --- | --- | --- | --- |
| Peak height [no. of deaths per 1 mln inhabitants] | 34 | 7.22 | 8.65 | 3.48 | 1.68 | 12.78 | 0.48 | 42.80 |
| Peak height [raw no. of deaths] | 34 | 180.50 | 322.67 | 32.50 | 8.00 | 185.00 | 2.00 | 1172.00 |
| Peak time [no. of days after first death reported] | 34 | 31.32 | 13.94 | 31.00 | 22.00 | 40.00 | 2.00 | 71.00 |
| All beds capacity [per 1 mln inhabitants] | 34 | 4940.25 | 1744.89 | 4634.17 | 3427.67 | 6623.84 | 2512.45 | 8248.80 |
| ICU beds capacity [per 1 mln inhabitants] | 34 | 141.14 | 82.61 | 110.94 | 86.50 | 191.75 | 20.72 | 349.20 |
| Total no. of tests up to time of the peak [per 1 mln inhabitants] | 34 | 763.09 | 743.98 | 526.54 | 443.77 | 824.78 | 148.92 | 4166.11 |
| Stay-at-home order day | 34 | 8.74 | 10.33 | 8.00 | 1.00 | 14.00 | -6.00 | 41.00 |
| Educational facilities closure day | 34 | -0.26 | 8.31 | 0.50 | -3.00 | 4.00 | -22.00 | 17.00 |
| Gathering restrictions day | 34 | -1.76 | 8.67 | -1.00 | -6.00 | 2.00 | -21.00 | 17.00 |
| Businesses closure day | 34 | 3.32 | 10.14 | 2.00 | -1.00 | 7.00 | -16.00 | 41.00 |
| Population size [Jan 2020] | 34 | 23.82 | 33.02 | 9.33 | 4.94 | 37.85 | 0.34 | 145.93 |
| Proportion living in urban areas | 34 | 0.74 | 0.12 | 0.74 | 0.66 | 0.83 | 0.54 | 0.98 |
| Proportion living in metropolitan cities with more than 1 mln inhabitants | 34 | 0.28 | 0.19 | 0.31 | 0.12 | 0.44 | 0.00 | 0.56 |
| Median age | 34 | 41.85 | 2.96 | 42.50 | 40.50 | 44.00 | 32.00 | 46.70 |
| Arrivals at airports in 2018 [per 1 inhabitant] | 34 | 2.27 | 2.63 | 1.52 | 0.84 | 2.79 | 0.24 | 14.86 |
| No. of foreign tourists in 2018 [per 1 inhabitant] | 34 | 1.25 | 1.32 | 0.79 | 0.47 | 1.46 | 0.14 | 6.73 |
| Mobility score at the day of first reported death | 34 | -23.45 | 18.93 | -20.02 | -44.09 | -5.28 | -56.42 | 0.16 |
| Border closure day | 34 | 6.71 | 15.53 | 2.50 | -1.00 | 11.00 | -19.00 | 46.00 |
| No. of COVID-19 infections when borders were closed [per 1 mln inhabitants] | 34 | 490.54 | 1116.56 | 83.39 | 21.53 | 245.88 | 0.07 | 5223.83 |
| No. of COVID-19 deaths when borders were closed [per 1 mln inhabitants] | 34 | 28.56 | 71.73 | 0.27 | 0.00 | 3.17 | 0.00 | 297.90 |

Abbreviations: ICU, intensive care unit; SD, standard deviation; mln, million.

**Figure 1.**
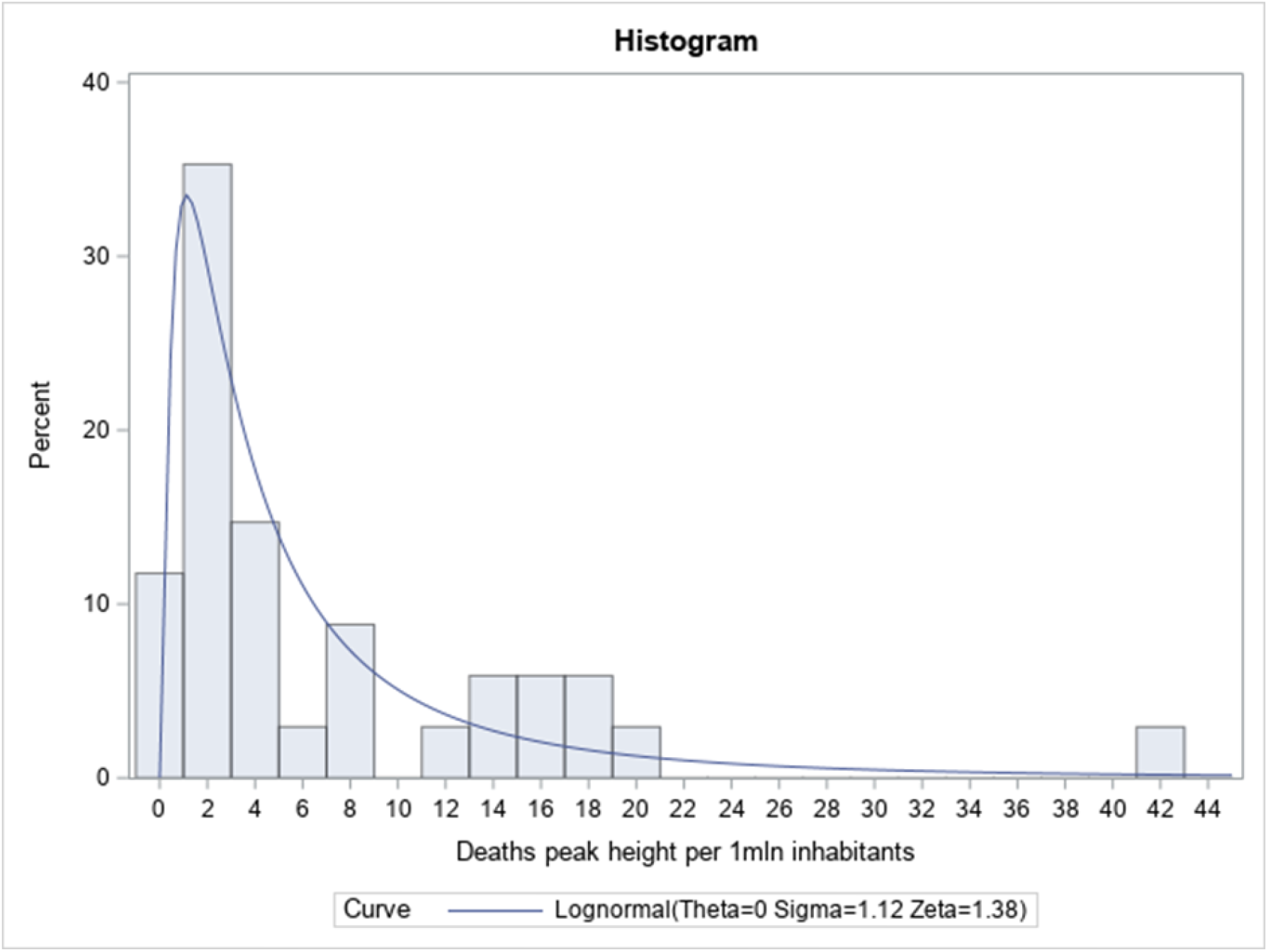
Histogram of the height of COVID-19 daily deaths peak per 1 million inhabitants with a fitted log-normal curve Histogram presents the distribution of height of COVID-19 deaths peak per 1 million inhabitants across 34 countries. A log-normal curve was fitted to the distribution with scale parameter sigma=1.12 and location parameter zeta=1.38, assuming logarithm of height of the deaths peak is normally distributed [Normal(zeta, sigma)].

**Figure 2.**
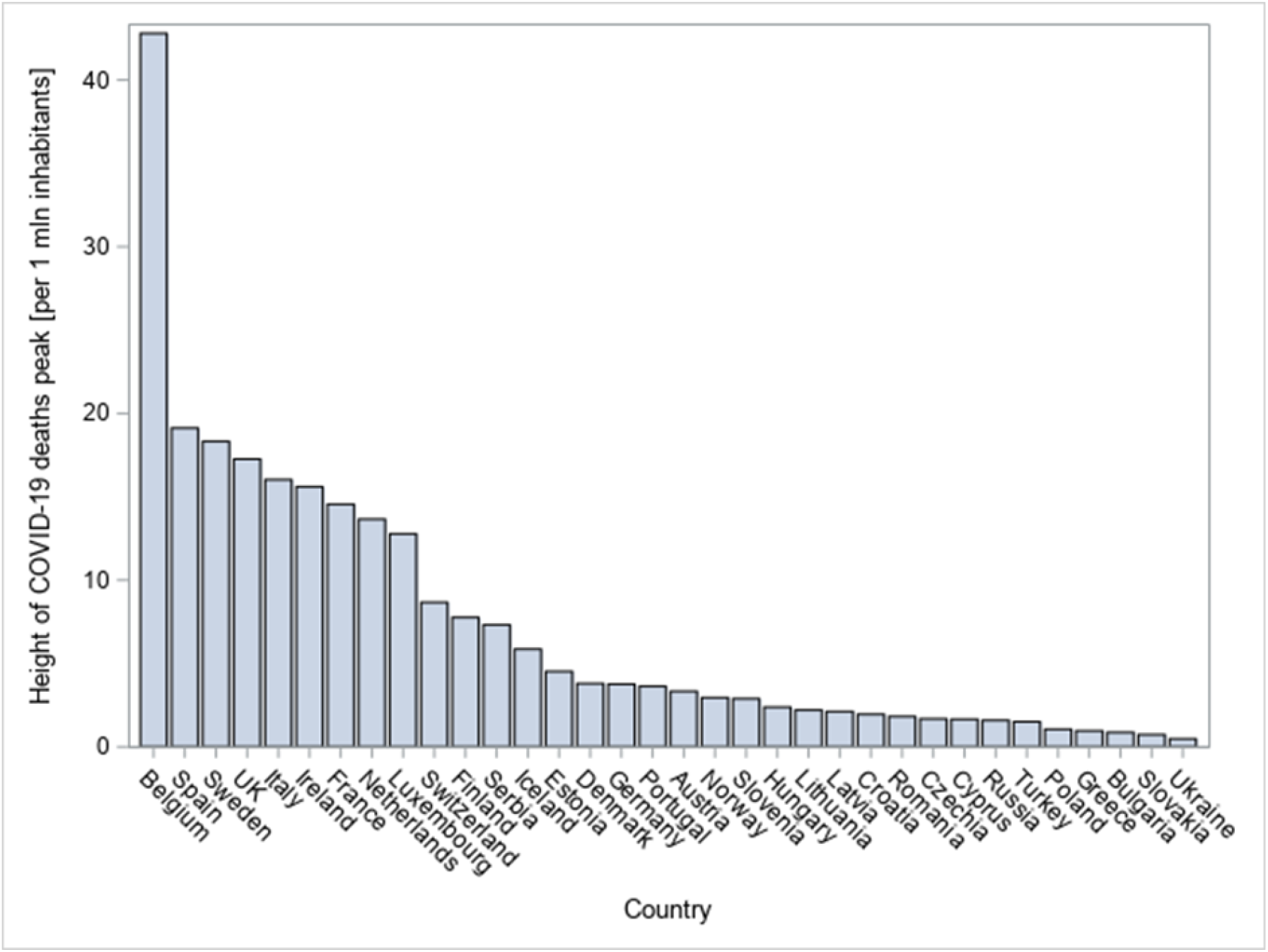
Height of COVID-19 daily deaths peak per 1 million inhabitants across countries The plot presents the height of COVID-19 deaths peak per 1 million inhabitants across 34 countries, with Belgium having outstandingly highest peak per population size (>40 deaths per 1 million inhabitants).

Median height of the peak per 1 million inhabitants was equal to 3.48 deaths per day ([lower quartile; upper quartile] = [1.68; 12.78]), with Belgium reaching outstandingly higher peak than other countries. As can be seen from these statistics and the histogram, the height of the deaths peak does not follow a normal distribution, but it can be well approximated using a log-normal distribution. Median time to the peak equalled 31 days from the first death reported in each country with a comparable mean (31.32, SD=13.94). Average numbers of days when closing schools/universities and when gatherings were banned were both negative (−0.26 and −1.76, respectively).

Moran’s I and Geary’s C statistics for height of the deaths peak were discordant, therefore spatial autocorrelation was not considered in the main analysis of deaths peak height, but considered in sensitivity analysis (Supplementary Materials). Moran’s I and Geary’s C statistics for time to peak both indicated no significant spatial autocorrelation, therefore this was not considered in the analysis of time to peak.

### Height of the deaths peak

Univariate GLM models of height of the peak were analysed (Table 2). Eight factors turned out to be significant: educational facilities closure day, gathering restrictions day, businesses closure day, proportion living in urban areas, mobility score at the day of first reported death, border closure day, number of COVID-19 infections when borders were closed, and number of COVID-19 deaths when borders were closed. All of them have positive estimates. Stay-at-home order day, as well as the proportion living in metropolitan cities >1 million inhabitants were close to reaching the significance level (p<0.1). Pearson correlations between variables reaching or close to reaching significance in univariate models were verified (Supplementary Materials).

**Table 2.** Results of univariate and multivariate GLM of COVID-19 deaths peak height, with normal distribution and logit link function

| Univariate analysis |  |  |  |  | Multivariate analysis |  |  |  |  |  |  |  |
| --- | --- | --- | --- | --- | --- | --- | --- | --- | --- | --- | --- | --- |
|  |  |  |  |  | Base case GLM<br>(Full model) |  |  |  | Base case GLM + selection algorithms<br>(Final model) |  |  |  |
| Variable | Estimate | Wald 95%<br>confidence limit |  | p-value | Estimate | Wald 95% confidence<br>limit |  | p-value | Estimate | Wald 95%<br>confidence limit |  | p-value |
|  |  | Lower | Upper |  |  | Lower | Upper |  |  | Lower | Upper |  |
| All beds capacity [per 1 mln inhabitants] | -0.0002 | -0.001 | 0.0001 | 0.109 | - | - | - | - | - | - | - | - |
| ICU beds capacity [per 1 mln inhabitants] | -0.002 | -0.008 | 0.003 | 0.404 | - | - | - | - | - | - | - | - |
| Total no. of tests up to time of the peak [per 1 mln inhabitants] | 0.000 | -0.001 | 0.001 | 0.961 | - | - | - | - | - | - | - | - |
| Stay-at-home order day | 0.027 | -0.001 | 0.053 | 0.057** | - | - | - | - | - | - | - | - |
| Educational facilities closure day | 0.067 | 0.028 | 0.107 | 0.001* | - | - | - | - | - | - | - | - |
| Gathering restrictions day | 0.045 | 0.006 | 0.084 | 0.023* | - | - | - | - | - | - | - | - |
| Businesses closure day | 0.029 | 0.009 | 0.049 | 0.005* | -0.001 | -0.022 | 0.020 | 0.944 | - | - | - | - |
| Population size [mln] | 0.001 | -0.009 | 0.011 | 0.877 | - | - | - | - | - | - | - | - |
| Proportion living in urban areas | 8.117 | 3.695 | 12.539 | <0.001* | 7.127 | 3.642 | 10.611 | <0.001* | 6.848 | 4.016 | 9.680 | <0.001* |
| Proportion living in metropolitan cities with more than 1 mln inhabitants | 2.428 | -0.464 | 5.319 | 0.099** | 1.063 | -1.331 | 3.457 | 0.384 | - | - | - | - |
| Median age | -0.0003 | -0.120 | 0.120 | 0.996 | - | - | - | - | - | - | - | - |
| Arrivals at airports in 2018 [per 1 inhabitant] | 0.016 | -0.097 | 0.129 | 0.784 | - | - | - | - | - | - | - | - |
| No. of foreign tourists in 2018 [per 1 inhabitant] | -0.076 | -0.421 | 0.270 | 0.668 | - | - | - | - | - | - | - | - |
| Mobility score at the day of first reported death | 0.048 | 0.001 | 0.096 | 0.046* | 0.041 | -0.0004 | 0.083 | 0.052* | 0.049 | 0.022 | 0.077 | <0.001* |
| Borders closure day | 0.025 | 0.010 | 0.041 | 0.002* | -0.005 | -0.030 | 0.021 | 0.729 | - | - | - | - |
| No. of COVID-19 infections when borders were closed [per 1 mln inhabitants] | 0.0002 | 0.0000 | 0.004 | 0.022* | 0.0003 | -0.0001 | 0.001 | 0.113 | 0.0002 | 0.0001 | 0.0003 | 0.016* |
| No. of COVID-19 deaths when borders were closed [per 1 mln inhabitants] | 0.004 | 0.001 | 0.007 | 0.002* | - | - | - | - | - | - | - | - |
| Multivariate models statistics |  |  |  |  |  |  |  |  |  |  |  |  |
| Scale | - | - | - | - | 5.020 | 3.958 | 6.367 | - | 5.092 | 4.015 | 6.458 | - |
| AIC | - | - | - | - | 222.203 | - | - | - | 217.169 | - | - | - |
| AICC | - | - | - | - | 227.963 | - | - | - | 219.312 | - | - | - |

\*p-value <0.05; \*\*p-value < 0.1.
Abbreviations: AIC, Akaike's Information Criterion; AICC, AIC corrected for small sample sizes; GLM, generalised linear models; ICU, intensive care unit; mln, million.
GLM univariate and multivariate models with normal distribution and logit link function were used to explore factors associated with COVID-19 deaths peak height as of 3<sup>rd</sup> June 2020. Each model was run using 34 observations. Variables significant in univariate models were included into the multivariate base case model, avoiding highly correlated pairs. The final multivariate model was selected based on the use of selection algorithms (backward, forward, stepwise and the genetic algorithm).

A base case multivariate GLM model included six covariates. The proportion living in urban areas was found significant (p<0.001) and the mobility score at the day of first reported death was very close to reaching significance (p=0.052). Results are presented in Table 2. Considering small sample size (N=34), selection algorithms were applied to the base case model. All algorithms were consistent and selected the model with three significant parameters (“final” model), indicating its best fit. The proportion of the population living in urban areas (6.848, p<0.001), mobility score at the day of first reported death (0.049, p<0.001) and number of infections when borders were closed per 1 million inhabitants (0.0002, p=0.016) were all significantly associated with the deaths peak height (Table 2).

Analysis of data removing countries with imputed dates of businesses or borders closure (N=29) provided consistent results regarding the significance of all three covariates from the final models (Supplementary Materials). In the model with spatial autocorrelation, the effects of the proportion of the population living in urban areas and of number of infections when borders were closed remained positive and statistically significant (p=0.022 and 0.012, respectively), but the effect of mobility score was no longer significant (p=0.400) (Supplementary Materials).

### Time to the deaths peak

Univariate GLM models of time to the peak were analysed (Table 3). Nine factors turned out to be potentially significant (with p<0.1): all beds capacity, ICU beds capacity, educational facilities closure day, gathering restrictions day, population size, arrivals at airports per inhabitant, number of foreign tourists per inhabitant, mobility score at the day of first reported death, and day of border closure. Most of these factors have positive estimates, except the numbers of arrivals and tourists. As for the next step, linear correlations between them were verified. Results can be found in Supplementary Materials.

**Table 3.**
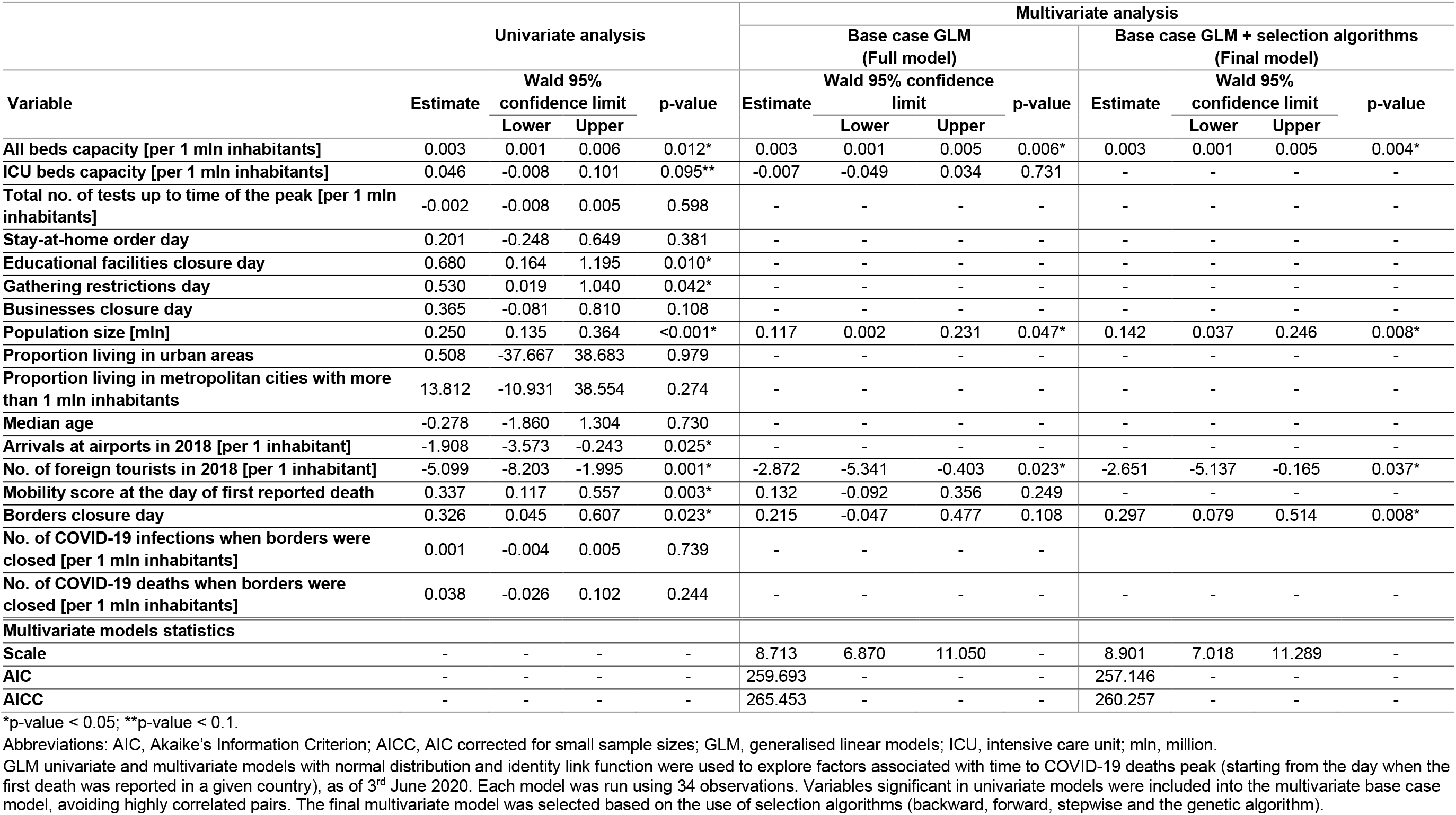
Results of univariate and multivariate GLM of time to COVID-19 deaths peak, with normal distribution and identity link function

A base case multivariate GLM model was run using six explanatory variables. All beds capacity, population size in millions and the number of foreign tourists per inhabitant were found significantly related with time to COVID-19 deaths peak. Results are presented in Table 3. Considering the small sample size (N=34), selection algorithms were applied to the above multivariate model. The “final” model had four covariates and was selected by the backward and the genetic algorithm (Supplementary Materials). All beds capacity per 1 million inhabitants (0.003, p=0.004), population size in millions (0.142, p=0.008), the number of foreign tourists per inhabitant (−2.651, p=0.037) and border closure day (0.297, p=0.008) were all significantly associated with time to the deaths peak (Table 3).

Analysis of data removing countries with imputed borders closure dates (N=31) provided consistent results regarding the significance of all beds capacity and population size, whereas number of foreign tourists and borders closure day were close in reaching significance, with p<0.1 (Supplementary Materials).

## Discussion

This study provides some evidence about factors associated with COVID-19 mortality peak and time to the peak. One of the strongest predictors of the COVID-19 mortality peak identified in our analysis was the proportion of population living in urban areas. The relationship between urbanisation and COVID-19 infections ratio was earlier outlined by the United Nations Association,^26^ estimating that 90% of all reported COVID-19 cases (by July 2020) came from urban areas, becoming the epicentre of the pandemic. High population density increases the propensity of viruses spread by increasing the contact rates of individuals. However, some authors^27^ warn readers against putting too much weight on urban density, arguing that large cities just faced the coronavirus earlier, due to the higher number of incoming people, and that the timing of epidemy start was of bigger interest than population density itself. This observation seems to be on the contrary to our study, since neither proportion of population living in urban areas, nor in metropolitan cities were related with time to deaths peak.

Number of infections when borders were closed was found to be another important factor associated with the COVID-19 deaths peak height, whereas a positive association between borders closure day and time to reach the peak was observed. It shows that stopping arrivals to the country at the earlier stage of epidemy can be crucial in reducing the peak height, stopping the increase of daily number of deaths earlier and, consequently, to flatten the deaths curve. These findings are on the contrary with Chaudhry who showed no association between rapid border closures and COVID-19 mortality using cumulative data as of April 2020.^28^ However, the peak height and mortality may not be strongly correlated. For example, some countries experienced a point peak and dropping fast (France, Spain, Italy), while other countries experienced a flattened peak with mortality staying high over a long period of time (US, Brazil, Mexico).^29^

The difference in average mobility before the pandemic and at the day when first death was reported in a country was found to be associated with deaths peak height. Previous studies confirm that changes in social-distancing covariates can flatten the curve by changing the peak death rate.^9, 10^ The impact of mobility was no longer significant when accounting for spatial autocorrelation, but this might be due to similarity in policies between neighbouring countries leading to similar outcomes.

Tourism indicators were found not to be associated with deaths peak height, but to be significant drivers of time to reach the peak. It suggests that the magnitude of inbound tourism can have an impact on accelerating the moment of reaching the peak. However, Aldibasi^5^ and Garcia de Alcaniz^8^ found tourism to be a significant predictor of COVID-19 mortality and infection rates. Ostig and Askin^30^ also found a significant positive relationship between number of airline passengers and number of COVID-19 infections.

Another conclusion resulting from the study is that timing of government restrictions, especially border closure, can be found as an important factor in terms of COVID-19 mortality. What can be observed for many countries is that government restrictions took place several days before any death was reported in a country. Therefore, increase or decrease in mortality can be interpreted as a result of undertaking preventive action by the government. However, when some countries began lockdown too late (Italy, France, United Kingdom), others may have started it earlier than necessary (Poland, Slovakia). Previous studies suggested that there are no significant benefits of very restrictive interventions such as mandatory stay-at-home order.^31, 32^ Furthermore, prevention and treatment of noncommunicable diseases have been severely disrupted since the pandemic began.^33, 34^ Thus, the balance between confinement timing and avoidance of prolonged lockdown is essential to prevent societies from deep recession^35^ and negative consequences on health outcomes.

There is no general conclusion on the association between COVID-19 fatality and hospital beds capacity in literature. Our study showed lack of association between beds capacity and deaths peak height, but higher beds capacity was related with longer time to peak. Garcia de Alcaniz showed no association between hospital beds density and both the number of infections and number of deaths at any moment of the pandemic.^8^ On the contrary, Sorci showed that case fatality rate was negatively associated with number of hospital beds per inhabitants.^7^ High bed capacity may be associated with more intensive treatment delaying time to death for a number of patients, thus delaying time to peak but does not affect the peak height.

## Limitations

Our study has several limitations. Data on a limited number of countries were used. We decided not to include countries for which comparative evidence could not be found. Our idea was to focus only on Europe, not to merge it with countries from other continents and avoid data incomparability issues and eventual difference in strain as the virus mutate rapidly.

However, number of observations can be assessed as high enough to draw reliable conclusions. Stability of results was tested with different selection algorithms, being consistent with main findings.

The set of included explanatory variables can be viewed as non-exhaustive. We decided to focus on variables important from a social perspective, assuming that urbanisation, mobility, tourism and government restrictions are more of interest to inform upcoming decisions than other approaches observed in the literature; for example, environmental perspective.^8, 11^

Another limitation was associated with data imputation. Countries with imputed government restriction dates were therefore excluded in sensitivity analysis, leading to generally consistent findings.

## Conclusions

This study demonstrated significant drivers of COVID-19 mortality magnitude in Europe and shed a light on reasons behind variability between countries. The higher proportion of population living in urban areas, lack of rapid reduction in mobility and delayed government restrictions correlated with a higher COVID-19 deaths peak. These findings can help improve future response in similar situations or in case of new waves.

## Data Availability

The authors confirm that the data supporting the findings of this study are available within the article and its supplementary materials.

## Acknowledgements

The authors wish to thank Saima Khan (Apothecom) for proof reading and editing support.

## Ethical approval

Not required. This study does not require ethical approval as it was conducted on country-level data and involved information freely available in the public domain.

## Contributors

MT conceived the study, KJ collected and analysed the data, KJ, MT and SA interpreted the results, KJ wrote the first draft of the manuscript, KJ, MT and SA revised the manuscript and approved the final version.

