## Supplementary Materials for "Factors influencing the COVID-19 daily deaths peak across European countries"

### Height of the COVID-19 deaths peak

#### Correlations between significant variables

Correlations between variables reaching or close to reaching significance in univariate models of the deaths peak height were verified. Results are presented in **Supplementary Table 1**. High correlations could be observed between educational facilities closure day and: gathering restrictions day (0.84, $p<0.001$), border closure day (0.76, $p<0.001$) and mobility score at the day of first reported death (0.76, $p<0.001$). The latter one was also highly correlated with gathering restrictions day (0.74, $p<0.001$). In addition, the number of deaths when closing borders highly correlated with border closure day (0.84, $p<0.001$). Correlations between number of deaths and number of infections when closing borders and between number of infections and border closure day were close to reaching the high level (0.69, $p<0.001$, both).

**Supplementary Table 1.** Pearson correlations between explanatory variables potentially significant (with p<0.1) in univariate GLM of deaths peak height

| | Pearson Correlation Coefficients | | | | | | | | | |
|---|---|---|---|---|---|---|---|---|---|---|
| | Educational facilities closure day | Gathering restrictions day | Sta-at-home order day | Businesses closure day | Proportion living in urban areas | Proportion living in metropolitan cities | Mobility score at the day of first reported death | Border closure day | No. of infections when borders were closed | No. of deaths when borders were closed |
| **Educational facilities closure day** | 1.00 | **0.84 p<0.001** | 0.31 p=0.07 | 0.51 p<0.001 | 0.41 p=0.02 | 0.34 p=0.05 | **0.76 p<0.001** | **0.76 p<0.001** | 0.29 p=0.09 | 0.52 p<0.001 |
| **Gathering restrictions day** | | 1.00 | 0.18 p=0.30 | 0.44 p=0.01 | 0.41 p=0.02 | 0.31 p=0.08 | **0.74 p<0.001** | 0.65 p<0.001 | 0.26 p=0.14 | 0.48 p<0.001 |
| **Sta-at-home order day** | | | 1.00 | 0.67 p<0.001 | 0.26 p=0.13 | 0.32 p=0.06 | 0.39 p=0.02 | 0.25 p=0.15 | 0.08 p=0.66 | 0.23 p=0.20 |
| **Businesses closure day** | | | | 1.00 | 0.36 p=0.04 | 0.48 p<0.01 | 0.58 p<0.001 | 0.38 p=0.02 | 0.17 p=0.34 | 0.31 p=0.08 |
| **Proportion living in urban areas** | | | | | 1.00 | 0.34 p=0.05 | 0.43 p=0.01 | 0.27 p=0.13 | 0.18 p=0.31 | 0.10 p=0.57 |
| **Proportion living in metropolitan cities** | | | | | | 1.00 | 0.39 p=0.02 | 0.31 p=0.07 | -0.03 p=0.86 | 0.26 p=0.14 |
| **Mobility score at the day of first reported death** | | | | | | | 1.00 | 0.62 p<0.001 | 0.21 p=0.23 | 0.36 p=0.04 |
| **Borders closure day** | | | | | | | | 1.00 | 0.69 p<0.001 | **0.84 p<0.001** |
| **Nb of infections when borders were** | | | | | | | | | 1.00 | 0.69 p<0.001 |
| **Nb of deaths when borders were** | | | | | | | | | | 1.00 |

GLM, generalised linear models; mln, million.

#### Sensitivity analysis – Selection algorithms

When applying stepwise, backward, forward and genetic selection algorithms to the base case model of deaths peak height, the model with four significant covariates was selected (**Supplementary Table 2**). The fact that the genetic algorithm selected this model indicates its best fit properties. The plot of AICC values across all possible models constructed with the use of covariates from the full model, is depicted on **Supplementary Figure 1**. The best model was associated with the lowest AICC value.

**Supplementary Table 2.** Results of multivariate GLM of COVID-19 deaths peak height when applying selection algorithms

| | Base case GLM + selection algorithms Final model | | | |
|---|---|---|---|---|
| **Variable** | **Estimate** | **Wald 95% confidence limit** | | **p-value** |
| | | **Lower** | **Upper** | |
| **Proportion living in urban areas** | 6.848 | 4.016 | 9.680 | <0.001* |
| **Mobility score at the day of first reported death** | 0.049 | 0.022 | 0.077 | <0.001* |
| **No. of COVID-19 infections when borders were closed [per 1 mln inhabitants]** | 0.0002 | 0.0001 | 0.0003 | 0.016* |
| **Scale** | 5.092 | 4.015 | 6.458 | - |
| **AIC** | 217.169 | - | - | - |
| **AICC** | 219.312 | - | - | - |

*p-value <0.05.
AIC, Akaike's Information Criterion; AICC, AIC corrected for small sample sizes; GLM, generalised linear models; mln, million.

**Supplementary Figure 1.** AICC values of models of height of deaths peak analysed with the use of genetic algorithm

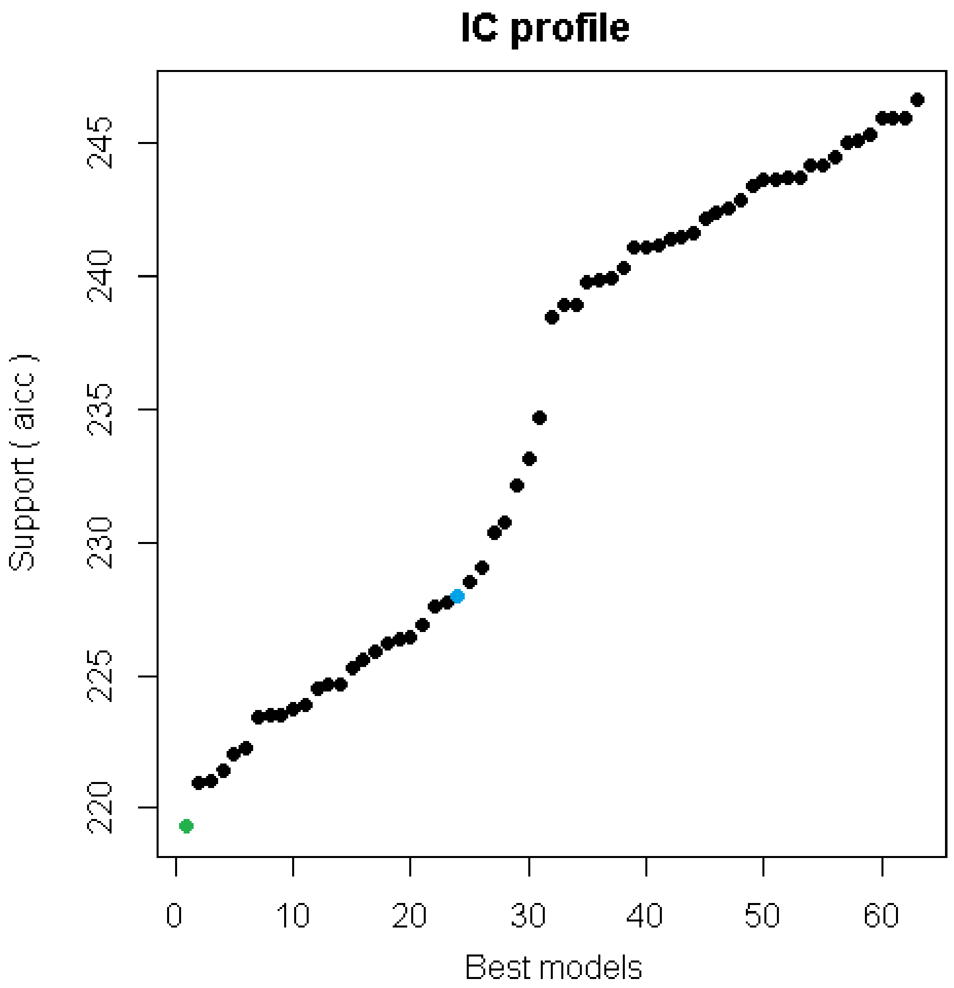


**Green dot** presents AICC of the model selected with all selection algorithms.
**Blue dot** presents AICC of the full model.
AICC, Akaike's Information Criterion corrected for small sample sizes.

#### Sensitivity analysis – Exclusion of countries with imputed government restriction dates

The base case and the final GLM models of deaths peak height were re-run on data after exclusion of countries for which government restriction dates, such as businesses or border closure dates, were imputed (N=29). Results are consistent with the base case findings in terms of significance of covariates of interest (**Supplementary Table 3**).

**Supplementary Table 3.** Results of multivariate GLM of COVID-19 deaths peak height excluding countries for which businesses or border closure dates were imputed (N=29)

| | Sensitivity analysis | | | | | | | |
|---|---|---|---|---|---|---|---|---|
| | SA of the full model | | | | SA of the final model | | | |
| Variable | Estimate | Wald 95% confidence limit | | p-value | Estimate | Wald 95% confidence limit | | p-value |
| | | Lower | Upper | | | Lower | Upper | |
| **Businesses closure day** | -0.118 | -0.275 | 0.040 | 0.143 | - | - | - | - |
| **Proportion living in urban areas** | 11.587 | 3.425 | 19.749 | 0.005* | 8.738 | 4.875 | 12.601 | <0.001* |
| **Proportion living in metropolitan cities with more than 1 mln inhabitants** | -1.457 | -4.616 | 1.702 | 0.366 | - | - | - | - |
| **Mobility score at the day of first reported death** | 0.347 | 0.104 | 0.591 | 0.005* | 0.064 | 0.026 | 0.103 | 0.001* |
| **Borders closure day** | -0.171 | -0.320 | -0.022 | 0.024* | - | - | - | - |
| **No. of COVID-19 infections when borders were closed [per 1 mln inhabitants]** | 0.007 | 0.003 | 0.012 | 0.001* | 0.001 | 0.0002 | 0.002 | 0.009* |
| **Scale** | 3.171 | 2.451 | 4.101 | - | 4.672 | 3.612 | 6.043 | - |
| **AIC** | 165.228 | - | - | - | 181.711 | - | - | - |
| **AICC** | 172.428 | - | - | - | 184.547 | - | - | - |

*p-value <0.05.
AIC, Akaike's Information Criterion; AICC, AIC corrected for small sample sizes; GLM, generalised linear models; mln, million; N, analysed sample size; SA, sensitivity analysis.

#### Sensitivity analysis – Spatial autocorrelation

Moran's I and Geary's C statistics were produced to check the existence of spatial autocorrelation in the values of deaths peak height across countries. The former reveals positive spatial autocorrelation, but the latter statistic does not confirm this finding (**Supplementary Table 4**).

**Supplementary Table 4.** Autocorrelation statistics under randomization assumption for values of height of the deaths peak across countries (N=34)

| Autocorrelation statistics | | | | | | |
|---|---|---|---|---|---|---|
| Variable | Assumption | Observed | Expected | Std dev | Z value | p-value |
| **Moran's I** | Randomization | 0.166 | -0.032 | 0.044 | 4.460 | <0.001* |
| **Geary's c** | Randomization | 0.830 | 1.000 | 0.238 | -0.715 | 0.474 |

*p-value <0.05

The final GLM model of the deaths peak height was then transformed into a spatial generalized linear mixed model (GLMM) to implement autoregressive structure. The estimated model is consistent with the base case analysis in terms of significance of covariates of interest, except mobility score which is no longer significant (p=0.40) (**Supplementary Table 5**).

**Supplementary Table 5.** Results of spatial GLMM of COVID-19 deaths peak height with implemented autoregressive structure (N=34)

| | Spatial GLMM model | | | |
|---|---|---|---|---|
| **Variable** | **Estimate** | **Wald 95% confidence limit** | | **p-value** |
| | | **Lower** | **Upper** | |
| **Proportion living in urban areas** | 3.305 | 0.514 | 6.095 | 0.022* |
| **Mobility score at the day of first reported death** | 0.008 | -0.011 | 0.026 | 0.400 |
| **No. of COVID-19 infections when borders were closed [per 1 mln inhabitants]** | 0.0004 | 0.0001 | 0.0010 | 0.012* |
| **AIC** | 192.640 | - | - | - |
| **AICC** | 195.750 | - | - | - |

*p-value <0.05.
AIC, Akaike's Information Criterion; AICC, AIC corrected for small sample sizes; GLM, generalised linear models; mln, million; N, analysed sample size.

### Time to the COVID-19 deaths peak

#### Distribution

**Supplementary Figure 2.** Time to COVID-19 daily deaths peak across countries

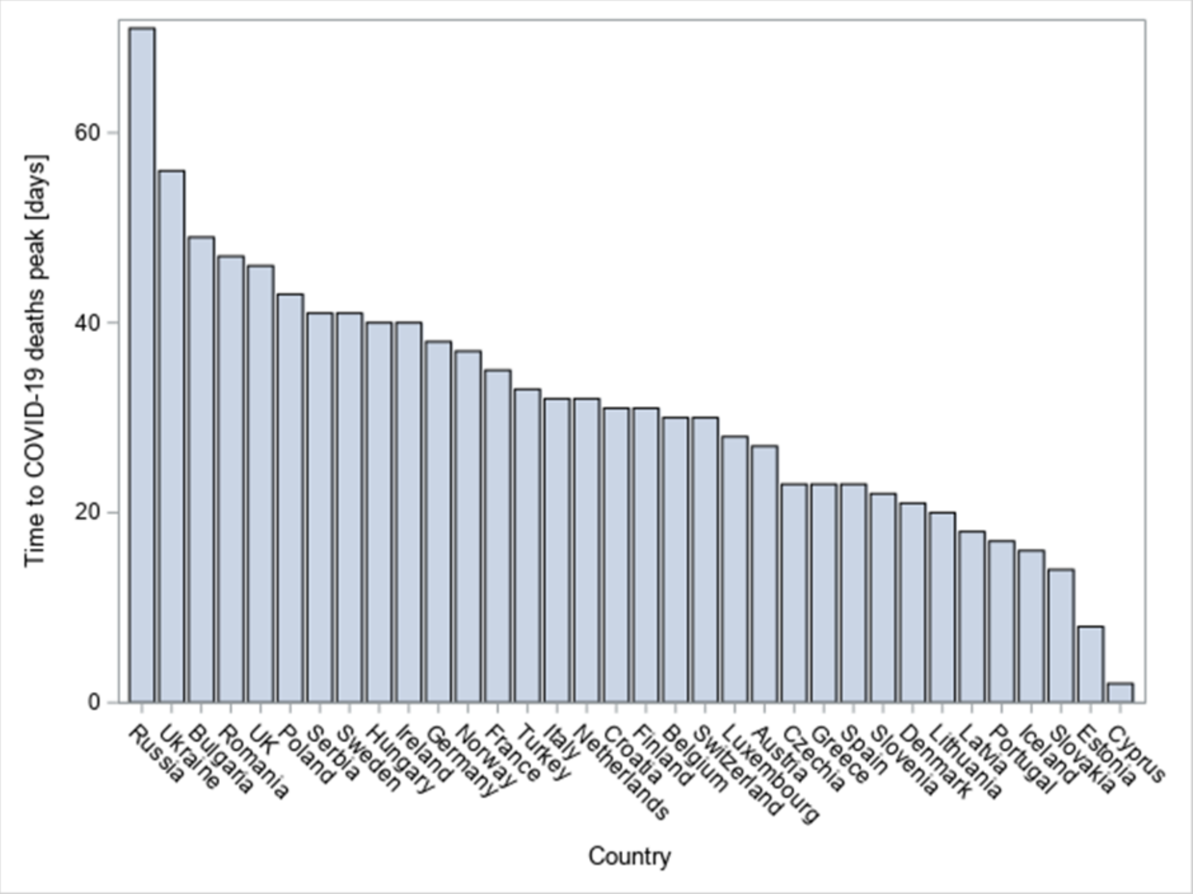

**Supplementary Figure 3.** Cumulative distribution of time to COVID-19 daily deaths peak

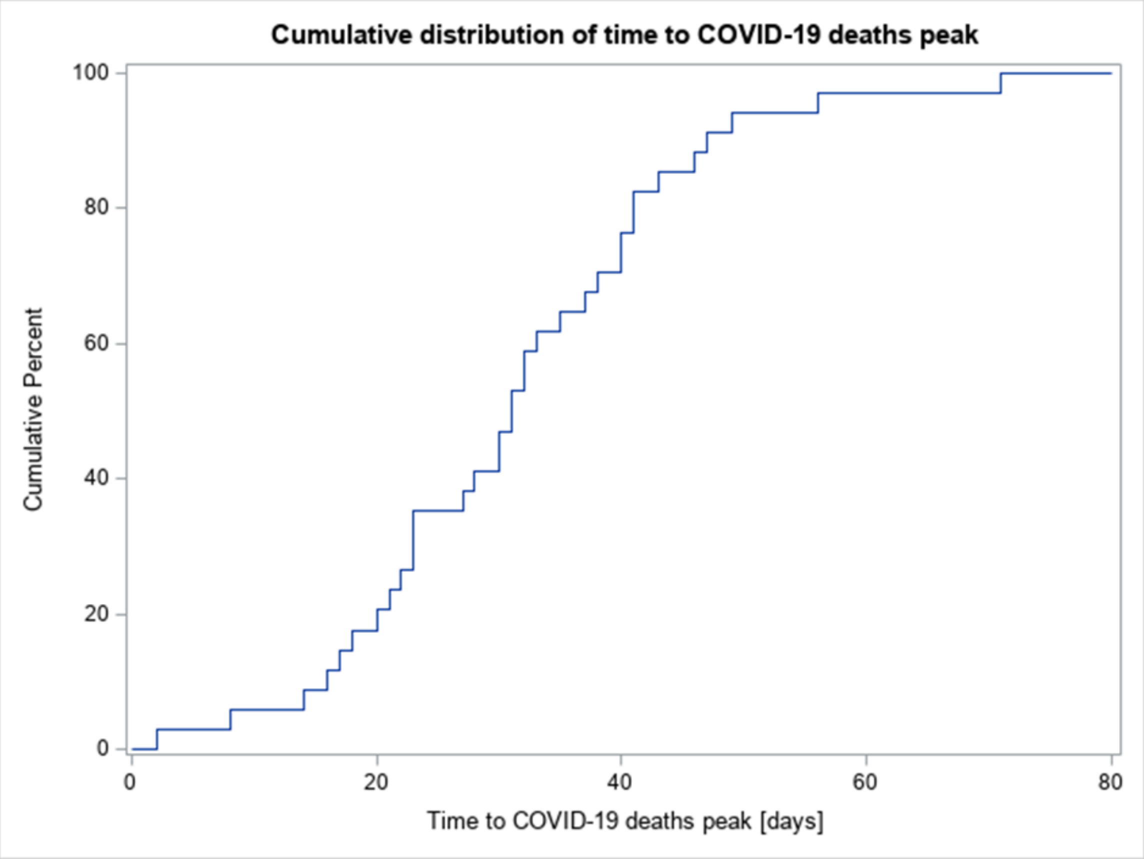


#### Correlations between significant variables

Correlations between variables reaching or close to reaching significance in univariate models of time to the peak were verified. Results are presented in **Supplementary Table 6**. High correlations could be observed between educational facilities closure day and: gathering restrictions day (0.84, p<0.001), border closure day (0.76, p<0.001) and mobility score at the day of first reported death (0.76, p<0.001). The latter one was also highly correlated with gathering restrictions day (0.74, p<0.001). The number of people that arrived at airports highly correlated with the number of foreign tourists (0.75, p<0.001).

**Supplementary Table 6.** Pearson correlations between explanatory variables potentially significant (with p<0.1) in univariate GLM models of time to deaths peak

| | Pearson Correlation Coefficients | | | | | | | | |
|---|---|---|---|---|---|---|---|---|---|
| | All beds capacity | ICU beds capacity | Educational facilities closure day | Gathering restrictions day | Population size [MLN] | Arrivals at airports | Number of foreign tourists | **Mobility score at the day of first reported death** | Border closure day |
| All beds capacity | 1.00 | 0.43 p=0.01 | -0.23 p=0.19 | -0.22 p=0.21 | 0.22 p=0.21 | -0.44 p=0.01 | -0.22 p=0.21 | -0.06 p=0.76 | -0.26 p=0.14 |
| ICU beds capacity | | 1.00 | 0.00 p=0.99 | 0.06 p=0.75 | 0.30 p=0.09 | -0.25 p=0.15 | -0.14 p=0.43 | 0.12 p=0.50 | 0.07 p=0.68 |
| Educational facilities closure day | | | 1.00 | **0.84 p<0.001** | 0.42 p=0.01 | 0.05 p=0.80 | -0.04 p=0.83 | **0.76 p<0.001** | **0.76 p<0.001** |
| Gathering restrictions day | | | | 1.00 | 0.56 p<0.001 | 0.14 p=0.44 | -0.05 p=0.78 | **0.74 p<0.001** | 0.65 p<0.001 |
| Population size [MLN] | | | | | 1.00 | -0.25 p=0.16 | -0.34 p=0.05 | 0.49 p<0.01 | 0.28 p=0.11 |
| Arrivals at airports | | | | | | 1.00 | **0.75 p<0.001** | 0.09 p=0.63 | 0.04 p=0.84 |
| Number of foreign tourists | | | | | | | 1.00 | -0.07 p=0.68 | -0.12 p=0.50 |
| **Mobility score at the day of first reported death** | | | | | | | | 1.00 | 0.62 p<0.001 |
| Border closure day | | | | | | | | | 1.00 |

GLM, generalised linear models; ICU, intensive care unit; mln, million.

#### Sensitivity analysis – Selection algorithms

When applying backward or genetic selection algorithms to the base case model of time to the peak, the model with four significant covariates was selected: all beds capacity, population size, number of foreign tourists and border closure day (**Supplementary Table 7**). The fact that the genetic algorithm selected the model indicates its best fit properties, hence this model was selected as the final one.

The stepwise and forward algorithms selected a similar model, but borders closure day was replaced by mobility score at the day of first reported death. However, fit statistics were slightly worse than those of the final selected model.

The plot of AICC values across all possible models constructed with the use of covariates from the full model, is depicted on **Supplementary Figure 4**. The best model was associated with the lowest AICC value.

**Supplementary Table 7.** Results of multivariate GLM of time to COVID-19 deaths peak when applying selection algorithms

| | Base case GLM + selection algorithms | | | | | | | |
|---|---|---|---|---|---|---|---|---|
| | Backward and genetic algorithms (Final model) | | | | Stepwise and forward algorithms | | | |
| Variable | Estimate | Wald 95% confidence limit | | p-value | Estimate | Wald 95% confidence limit | | p-value |
| | | Upper | Upper | | | Lower | Upper | |
| **All beds capacity [per 1 mln inhabitants]** | 0.003 | 0.001 | 0.005 | 0.004* | 0.002 | 0.0004 | 0.004 | 0.016* |
| **Population size [mln]** | 0.142 | 0.037 | 0.246 | 0.008* | 0.113 | -0.005 | 0.230 | 0.060** |
| **No. of foreign tourists in 2018 [per 1 mln inhabitants]** | -2.651 | -5.137 | -0.165 | 0.037* | -3.218 | -5.742 | -0.693 | 0.013* |
| **Mobility score at the day of first reported death** | - | - | - | - | 0.237 | 0.046 | 0.427 | 0.015* |
| **Border closure day** | 0.297 | 0.079 | 0.514 | 0.008* | - | - | - | - |
| **Scale** | 8.901 | 7.018 | 11.289 | - | 9.040 | 7.127 | 11.465 | - |
| **AIC** | 257.146 | - | - | - | 258.198 | - | - | - |
| **AICC** | 260.257 | - | - | - | 261.309 | - | - | - |

*p-value <0.05.
AIC, Akaike's Information Criterion; AICC, AIC corrected for small sample sizes; GLM, generalised linear models; mln, million.

**Supplementary Figure 4.** AICC values of models of time to deaths peak analysed with the use of genetic algorithm

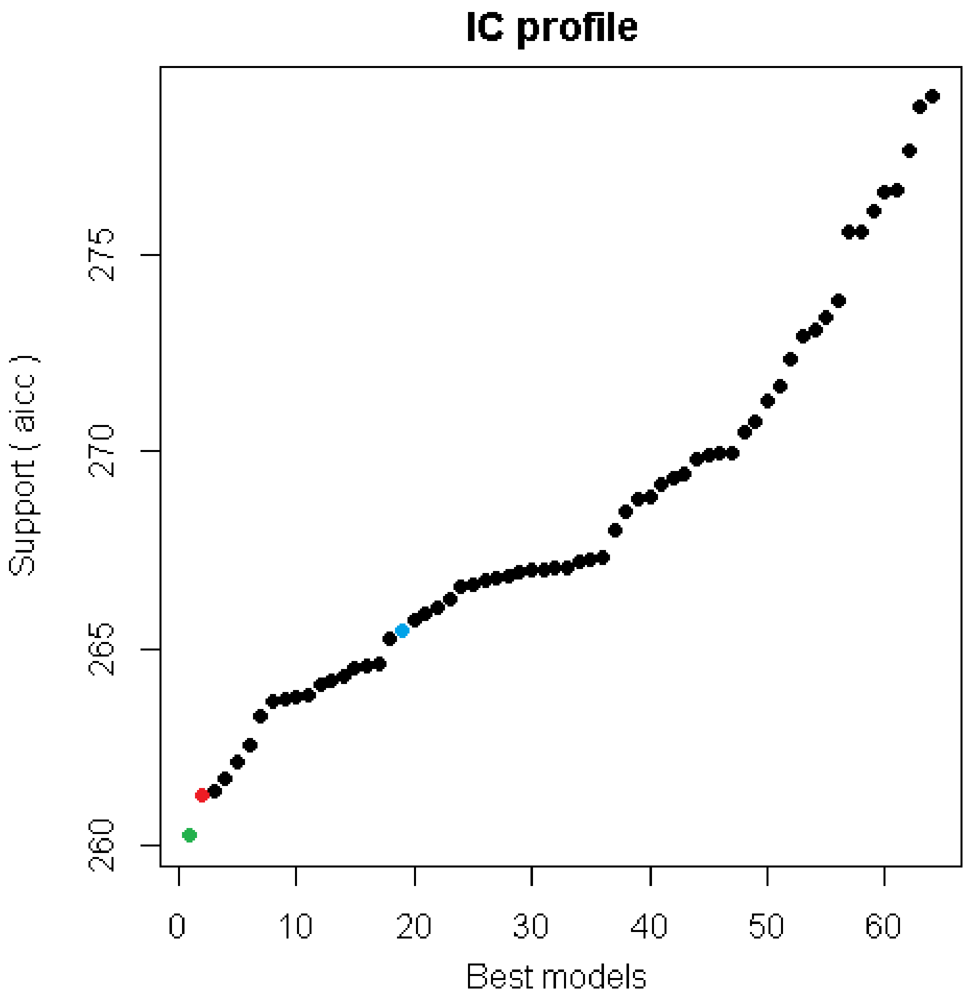


Green dot presents AICC of the model selected with backward and genetic selection algorithms.
Red dot presents AICC of the model selected with forward and stepwise selection algorithms.
Blue dot presents AICC of the full model.
AICC, Akaike's Information Criterion corrected for small sample sizes.

#### Sensitivity analysis – Exclusion of countries with imputed government restriction dates

The base case and the final GLM models of time to peak were re-run on data after excluding countries for which border closure dates were imputed (N=31). As a result, all beds capacity and population size remained significant in the final model, whereas remaining two variables, number of foreign tourists and border closure day, were close to reaching the significance level ($p<0.1$) (**Supplementary Table 8**).

**Supplementary Table 8.** Results of multivariate GLM of time to COVID-19 deaths peak excluding countries for which border closure dates were imputed (N=31)

| | Sensitivity analysis | | | | | | | |
|---|---|---|---|---|---|---|---|---|
| | SA of the full model | | | | SA of the final model | | | |
| Variable | Estimate | Wald 95% confidence limit | | p-value | Estimate | Wald 95% confidence limit | | p-value |
| | | Lower | Upper | | | Lower | Upper | |
| **All beds capacity [per 1 mln inhabitants]** | 0.003 | 0.001 | 0.005 | 0.008* | 0.003 | 0.001 | 0.005 | 0.005* |
| **ICU beds capacity [per 1 mln inhabitants]** | -0.003 | -0.048 | 0.042 | 0.904 | - | - | - | - |
| **Population size [mln]** | 0.117 | -0.008 | 0.242 | 0.067** | 0.143 | 0.026 | 0.259 | 0.017* |
| **No. of foreign tourists in 2018 [per 1 mln inhabitants]** | -2.755 | -5.341 | -0.169 | 0.037* | -2.529 | -5.144 | 0.085 | 0.058** |
| **Mobility score at the day of first reported death** | 0.143 | -0.100 | 0.386 | 0.248 | - | - | - | - |
| **Border closure day** | 0.151 | -0.221 | 0.524 | 0.426** | 0.273 | -0.029 | 0.576 | 0.077** |
| **Scale** | 9.051 | 7.057 | 11.609 | - | 9.250 | 7.211 | 11.864 | - |
| **AIC** | 240.553 | - | - | - | 237.898 | - | - | - |
| **AICC** | 247.098 | - | - | - | 241.398 | - | - | - |

*p-value <0.05; **p-value < 0.1.
AIC, Akaike's Information Criterion; AICC, AIC corrected for small sample sizes; GLM, generalised linear models; ICU, intensive care unit; mln, million; N, analysed sample size; SA, sensitivity analysis.

### Sensitivity analysis – Spatial autocorrelation

Moran's I and Geary's C statistics were produced to check the existence of spatial autocorrelation in the values of time to the deaths peak across countries. Both statistics do not reveal existence of spatial autocorrelation (**Supplementary Table 9**).

**Supplementary Table 9.** Autocorrelation statistics under randomization assumption for values of time to the deaths peak across countries (N=34)

| Autocorrelation statistics | | | | | | |
|---|---|---|---|---|---|---|
| Variable | Assumption | Observed | Expected | Std dev | Z value | p-value |
| **Moran's I** | Randomization | -0.030 | -0.032 | 0.048 | 0.047 | 0.963 |
| **Geary's c** | Randomization | 0.854 | 1.000 | 0.121 | -1.214 | 0.225 |
